# Statements issued by academic medical institutions after George Floyd’s killing by police and subsequent unrest in the United States: cross-sectional study

**DOI:** 10.1101/2020.06.22.20137844

**Authors:** Mathew V. Kiang, Alexander C. Tsai

## Abstract

**Background:** The horrific nature of George Floyd’s killing by a Minneapolis Police Department officer on May 25, 2020 has sparked more than a month of nationwide protests against police brutality and in support of the Black Lives Matter movement. At critical junctures of the nation’s public health such as these, academic medical institutions may exert leadership by issuing public statements to communicate institutional values.

**Methods:** We obtained statements issued by 56 leading U.S. medical schools relevant to George Floyd’s killing and subsequent protests. We tokenized statements into words, n-grams of sizes 2 and 3, and sentences; removed non-informative stop words and words that would compromise de-identification; and stemmed the remaining words using the Porter algorithm. We followed a predefined set of rules for identifying important elements of these statements related to leadership in antiracism and public health.

**Results:** Nearly all named George Floyd (50 [89%]), a majority noted the role of racism (43 [77%]) and acknowledged the Black community specifically (41 [73%]). Fewer ╌ slightly more than half ╌ referenced the act resulting in Floyd’s death (31 [55%]) or made explicit reference to the police (29 [52%]). Only 7 (13%) explicitly used terms denoting active support, like “antiracism” or “Black Lives Matter.” Most (45 [80%]) included references to negative sequelae resulting from racism like “disparities” or “inequality”. All included hopeful language.

**Conclusion:** Only a minority of institutions made reference to the killing of George Floyd by the police, and most failed to address this country’s targeted, historically engrained, and sustained oppression of Black people through white supremacy. Thus, our study identifies significant opportunities for U.S. medical schools to exert meaningful leadership in health.

## Introduction

On May 25, 2020, a Minneapolis Police Department officer killed George Floyd, a 46 year-old Black man, by kneeling on Floyd’s neck for 7 minutes and 46 seconds while he was handcuffed, lying prone, and pleading for his life. The horrific nature of the killing ╌ occurring in the context of longstanding disparities in police violence against Black people [1] and an unsuppressed national epidemic of coronavirus disease 2019 that has disproportionately burdened Black, Latinx, and American Indian communities [2] ╌ has sparked more than a month of nationwide protests against police brutality and in support of the Black Lives Matter movement. At critical junctures of the nation’s public health, academic medical institutions may exert leadership by issuing public statements to communicate institutional values; and enhance morale among faculty, staff, and students seeking reassurance of their value and who may be experiencing emotional distress [3]. The objective of this study was to describe statements issued by leading U.S. medical schools following George Floyd’s killing and subsequent protests.

## Methods

We obtained statements issued by 56 leading U.S. medical schools relevant to George Floyd’s killing and subsequent protests (**Supplement**; see also the publicly available Github repository corresponding to this analysis: https://github.com/mkiang/statement_analysis). After pre-processing the statements, we identified important elements each statement should contain based on defined criteria (**Supplement Table**): (1) use of victims’ names (e.g., “George Floyd”); (2) reference to Black people; (3) reference to the police; (4) specifies the act resulting in Floyd’s death; (5) explicitly naming racism (i.e., not “racial prejudice”); (6) explicitly using the term “antiracism”; (7) reference to negative sequelae resulting from racism (e.g., “inequity”); and (8) use of hopeful language (e.g., “community”, “inclusion”).

## Results

The median statement was issued on June 1, 2020 (interquartile range [IQR], May 31-June 2; range, May 29-June 2) and was 464 words in length (IQR, 371-560). Only three statements (5%) contained all of the defined elements (**Figure**). Nearly all named George Floyd (50 [89%]); fewer named either Breonna Taylor (23 [41%]) or Ahmaud Arbery (26 [46%]). A majority of statements noted the role of racism (43 [77%]) and acknowledged the Black community specifically (41 [73%]). Fewer ╌ slightly more than half ╌ referenced the act resulting in Floyd’s death (31 [55%]) or made explicit reference to the police (29 [52%]). Only 7 (13%) explicitly used terms denoting active support, like “antiracism” or “Black Lives Matter.” Most (45 [80%]) included references to negative sequelae resulting from racism like “disparities” or “inequality”. All included hopeful language.

**Figure.**
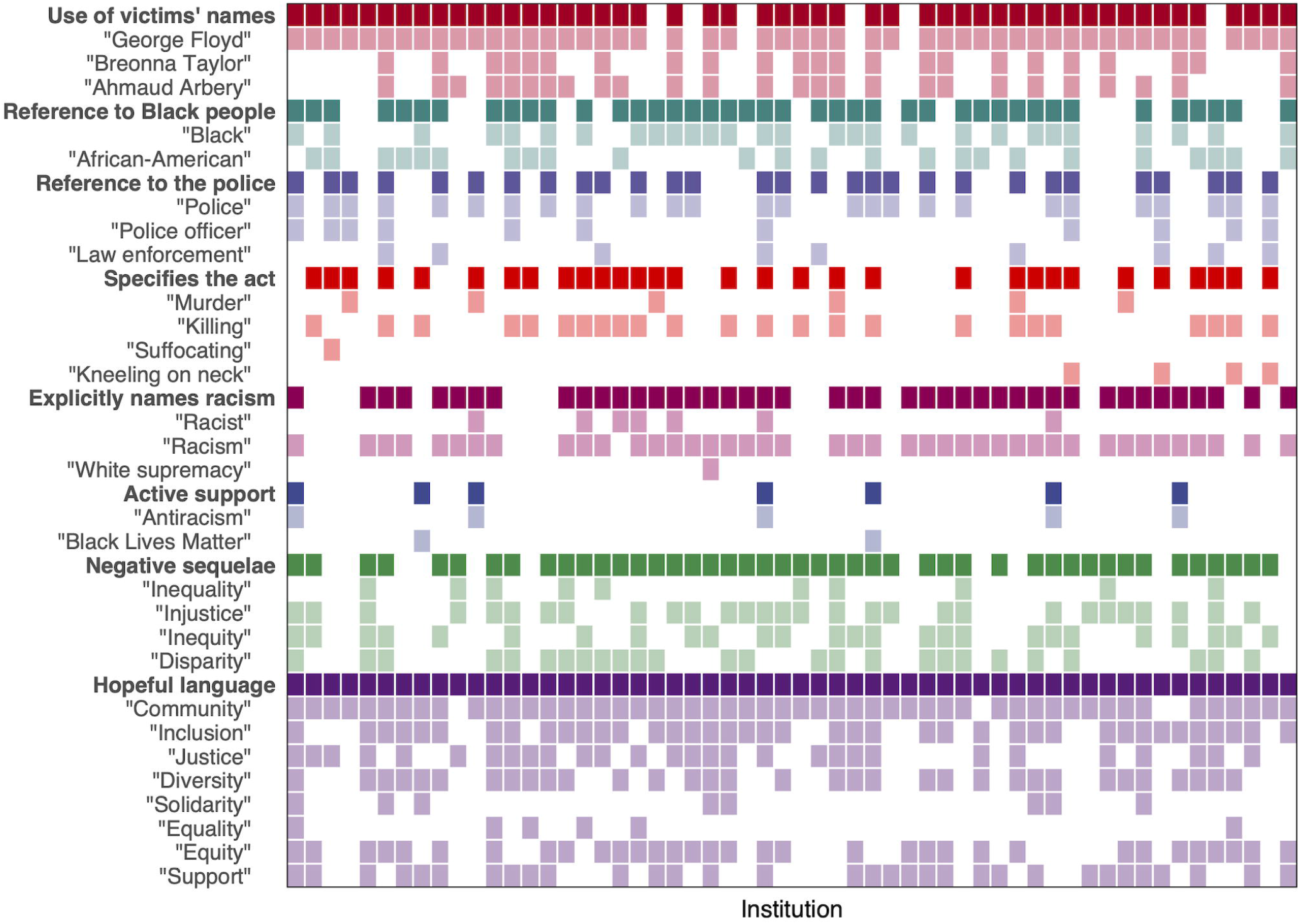
Presence of important elements of statements issued after George Floyd’s killing by police and subsequent unrest in the United States (y-axis), by medical school (x-axis) (n=56).

## Discussion

In this cross-sectional study of public statements issued by leading U.S. medical schools following the killing of George Floyd and subsequent protests, we found that only a minority of institutions made reference to the killing of George Floyd by the police, and most failed to address this country’s targeted, historically engrained, and sustained oppression of Black people through white supremacy. Thus, our study identifies significant opportunities for U.S. medical schools to exert meaningful leadership in health.

Two limitations likely bias our “quality assessments” toward the null. First, our analysis is limited to medical schools labeled as leading institutions according to ranking systems that are widely used but largely bereft of meaning [4,5]. Second, most academic medical institutions have failed to elevate Black people to the highest levels of institutional leadership [6].

Despite these limitations, our findings illustrate the need for leading academic medical institutions to lead amidst a chronic public health crisis disproportionately affecting Black people [1,3] layered upon an acute public health crisis disproportionately affecting Black, Latinx, and American Indian communities [2]. Possibilities include: articulating a clear stance on racism to affirm Black students, faculty, and staff and to motivate all students, faculty, and staff [3]; investing in financially costly initiatives to increase the proportion of community college and first-generation students, to increase the proportion of Black faculty members in leadership, to narrow Black/white disparities in faculty pay and tenure [4], and to ensure a living wage for full-time and subcontracted staff; training students in ways that do not contribute to the persistence of racism as a fundamental cause of Black/white health disparities; reviewing campus police practices to address racial profiling of Black students and faculty (however few there may be) and staff [1,3]; or committing to financially costless initiatives such as the renovation of educational spaces that venerate the white men who have historically dominated the field.

## Supplement

Kiang MV, Tsai AC. Statements issued by academic medical institutions after George Floyd’s killing by police and subsequent unrest in the United States: cross-sectional study.

### List of medical schools included in our analyses

We obtained statements relevant to George Floyd’s killing by police, and/or the subsequent protests, issued by U.S. medical schools identified in the top 50 ranking by either *U*.*S. News and World Report* list of “2021 Best Medical Schools: Research” or the 2018 total dollar amount of research awards from the U.S. National Institutes of Health. Due to ties in the rankings, the total sample includes statements from 56 medical schools:

| Institution | NIH Funding Rank<br>(2018) | USN&WR Rank<br>(2021) |
| --- | --- | --- |
| Albert Einstein College of Medicine | 32 | 40 |
| Baylor College of Medicine | 20 | 22 |
| Boston University | 45 | 29 |
| Brown University (Alpert) | 66 | 38 |
| Case Western Reserve University | 31 | 24 |
| Cleveland Clinic Lerner COM CWRU | 50 | NA |
| Columbia University | 8 | 6 |
| Cornell University (Weill) | 33 | 11 |
| Dartmouth College (Geisel) | 57 | 50 |
| Duke University | 9 | 12 |
| Emory University | 18 | 24 |
| Georgetown University | 78 | 44 |
| Harvard University | 26 | 1 |
| Icahn School of Medicine at Mount Sinai | 13 | 20 |
| Indiana University Indianapolis | 36 | 47 |
| Johns Hopkins University | 2 | 2 |
| Mayo Clinic School of Medicine | 22 | 6 |
| Medical College of Wisconsin | 49 | NA |
| Medical University of South Carolina | 43 | 58 |
| New York University (Grossman) | 19 | 4 |
| Northwestern University (Feinberg) | 16 | 18 |
| Ohio State University | 48 | 34 |
| Oregon Health and Science University | 25 | 28 |
| Stanford University | 3 | 4 |
| University of Michigan Ann Arbor | 10 | 15 |
| University of Alabama Birmingham | 21 | 31 |
| University of California Davis | 29 | 40 |
| University of California Irvine | 56 | 44 |
| University of California Los Angeles (Geffen) | 14 | 6 |
| University of California San Diego | 12 | 21 |
| University of California San Francisco | 1 | 6 |
| University of Chicago (Pritzker) | 38 | 17 |
| University of Cincinnati | 62 | 44 |
| University of Colorado | 23 | 31 |
| University of Florida | 46 | 40 |
| University of Iowa (Carver) | 41 | 34 |
| University of Kentucky | 47 | 68 |
| University of Maryland | 35 | 34 |
| University of Massachusetts Worcester | 34 | 47 |
| University of Miami (Miller) | 40 | 50 |
| University of Minnesota | 30 | 40 |
| University of North Carolina Chapel Hill | 17 | 23 |
| University of Pennsylvania (Perelman) | 5 | 3 |
| University of Pittsburgh | 6 | 14 |
| University of Rochester | 39 | 34 |
| University of South Florida | 85 | 47 |
| University of Southern California (Keck) | 27 | 31 |
| University of Texas Southwestern Medical Center | 28 | 26 |
| University of Utah | 37 | 38 |
| University of Virginia | 42 | 29 |
| University of Washington | 15 | 13 |
| University of Wisconsin Madison | 24 | 27 |
| Vanderbilt University | 11 | 18 |
| Wake Forest University Health Sciences | 44 | 52 |
| Washington University in St. Louis | 4 | 6 |
| Yale University | 7 | 15 |

### Additional information regarding methods

Given the focus of our analysis on academic medicine, we specifically identified statements made by the medical school dean. If multiple statements were issued, we analyzed the first statement issued. At institutions where a statement by the medical school dean could not be obtained, we substituted the statement made by someone at the institution’s next highest level of leadership (e.g., President or Chancellor).

We trimmed the statements to include only the full text of the statements, i.e., excluding the salutation (e.g., “Dear community”), signature (e.g., “Chief Diversity and Inclusion Officer”), and any specific resources identified in the statement (e.g., “*How to be an Antiracist*, by Ibram X. Kendi”). For one institution, the statement about George Floyd was embedded in a weekly newsletter and clearly identified as a discrete block of text. Following standard practice, we tokenized statements into words, n-grams of sizes 2 and 3, and sentences. We then removed non-informative stop words and words that would compromise de-identification (e.g., “Harvard” or “Stanford”). We stemmed the remaining words using the Porter algorithm. Stemming reduces words to their roots such as “communi” representing “community” and “communities”. Frequency tabulations of words and their stems, bigrams, and trigrams, both by institution and across all institutions, are available in the publicly available Github repository corresponding to this analysis (https://github.com/mkiang/statement_analysis).

After the preprocessing described above, we followed a predefined set of rules (**Table**) for identifying important elements of these statements related to leadership in antiracism and public health. These rules are described in both English and code in the Github repository (https://github.com/mkiang/statement_analysis).

**Table.**
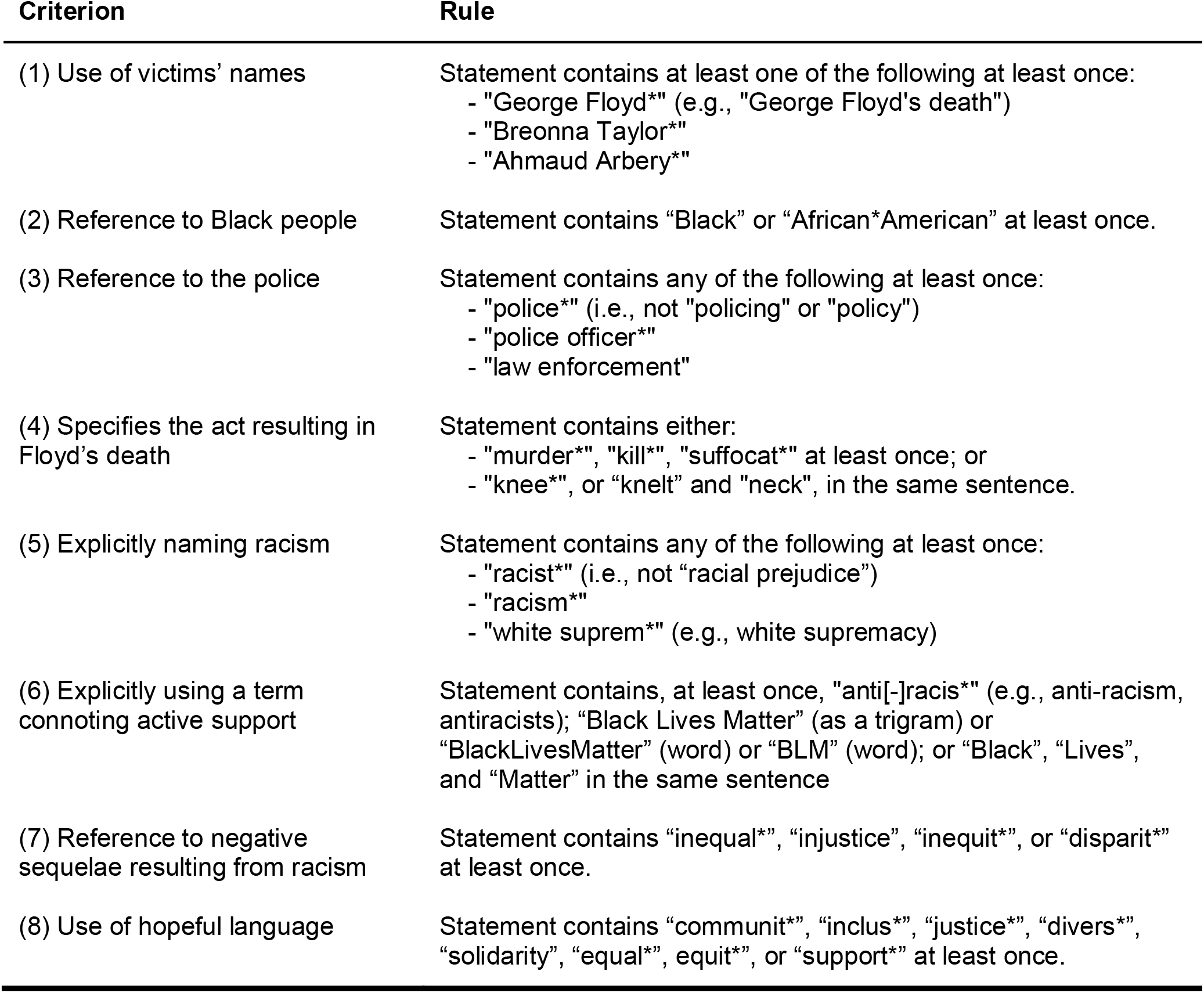
Criteria for identifying important elements of statements issued by leading medical schools after George Floyd’s killing and subsequent unrest in the United States

## Data Availability

Analysis code is available in the publicly available Github repository corresponding to this analysis.

https://github.com/mkiang/statement_analysis

